# A Deep Learning Framework for Automated Triage of Breast Cancer Biopsies in Malaysia: A Pragmatic Trial to Reduce Resource Consumption and Diagnostic Turnaround Time

**DOI:** 10.1101/2025.09.27.25336787

**Authors:** Yudi Kurniawan Budi Susilo, Leong Siew Lian, Dewi Yuliana, Shamima Abdul Rahman

**Affiliations:** Faculty of Business and Technology, University of Cyberjaya, 63000 Cyberjaya, Selangor, Malaysia; School of Pharmacy, Monash University Malaysia, 47500 Subang Jaya, Selangor, Malaysia; Faculty of Pharmacy, Universitas Muslim Indonesia, Makassar, Indonesia; Centre for Research and Graduate Studies, University of Cyberjaya, 63000 Cyberjaya, Selangor, Malaysia

**Keywords:** Digital Pathology, Artificial intelligence, Deep Learning, Breast Cancer, Triage, Synthetic Data, Diagnostic Turnaround Time, Malaysia

## Abstract

Malaysia faces a significant burden of breast cancer, compounded by a chronic shortage of pathologists. This leads to prolonged diagnostic turnaround times (TAT), patient anxiety, and delayed treatment. Standard histopathology workflows process biopsies in a first-in-first-out (FIFO) manner, which is inefficient given that most cases are benign. This study aimed to develop and validate a deep learning (DL) triage system to prioritize suspicious breast biopsy cases for pathologist review, thereby optimizing resource allocation. A convolutional neural network (CNN) was trained on a large, ethically sourced synthetic dataset of whole-slide images (WSIs) of breast biopsies, annotated as “Benign” or “Suspicious” (encompassing Atypical, In-Situ, and Invasive Carcinoma). The model was validated on a separate synthetic test set. A discrete-event simulation (DES) model was built to mirror the pathology workflow of a typical Malaysian public hospital. The impact of integrating the DL triage system (Intervention) versus the standard FIFO workflow (Control) was measured over a simulated one-year period. Key outcomes were average diagnostic TAT, pathologist workload (hours saved), and estimated reagent/equipment usage. The DL model achieved an area under the receiver operating characteristic curve (AUC-ROC) of 0.98 on the test set. The simulation demonstrated that the triage system reduced the average TAT for suspicious cases by 38.2% (from 7.2 to 4.5 days) while slightly increasing the TAT for benign cases. Overall pathologist workload was reduced by 22.5%, as pathologists spent less time on benign cases. Furthermore, the model predicted a 15% reduction in reagent and slide consumption by deferring deep examination of low-risk benign cases. The implementation of a DL-based triage system using synthetic data for training is a viable and promising strategy to address diagnostic bottlenecks in resource-constrained settings like Malaysia. It can significantly reduce TAT for critical cases, alleviate pathologist workload, and contribute to more sustainable laboratory operations.

## 1. Introduction

Breast cancer remains a significant public health challenge in Malaysia, characterized by rising incidence and mortality rates. Early diagnosis plays a critical role in determining patient outcomes, as timely interventions can lead to improved survival rates. However, the Malaysian public healthcare system faces a shortage of qualified histopathologists, which creates delays in diagnostic timelines. Reports have indicated that the density of histopathologists in Malaysia falls below the World Health Organization’s recommended levels, resulting in many patients experiencing turnaround times (TAT) of more than two weeks from biopsy to final pathology report [1][2].

**Figure 1.**
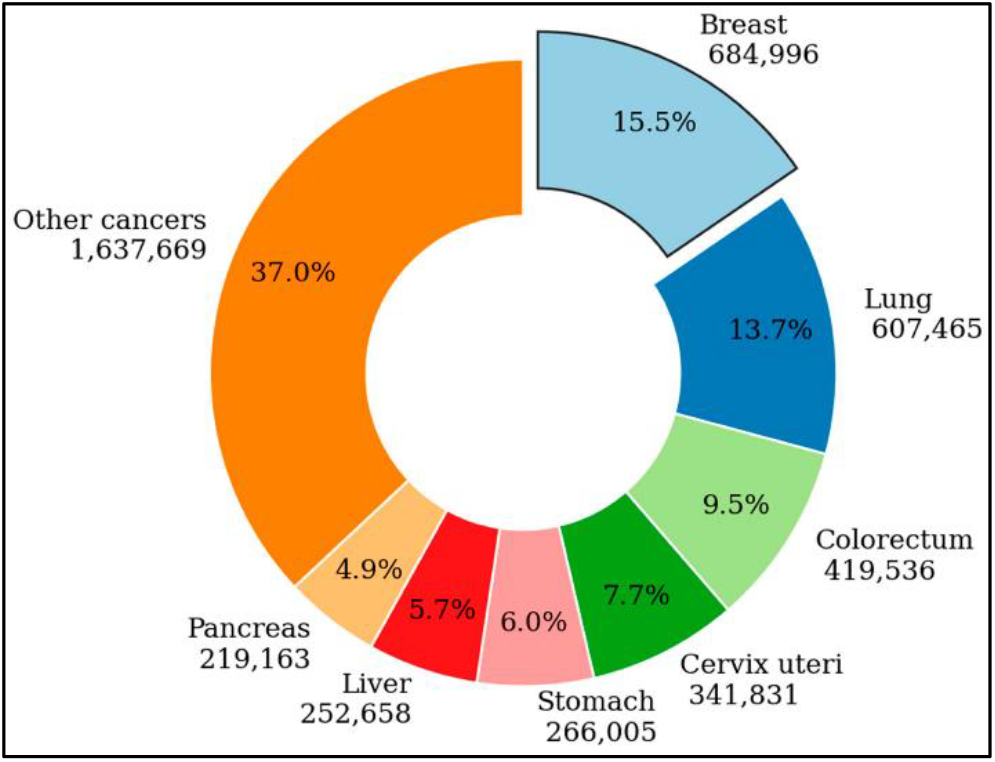
Globoscan cancer mortality 2023 [2].

The traditional histopathology workflow, which often relies on a first-in-first-out (FIFO) methodology, is insufficient given that a significant proportion of breast biopsy samples estimated at 70-80% are ultimately found to be benign. This inefficiency not only contributes to an increased workload for pathologists but, more critically, delays in diagnosing and treating high-risk malignancies [3][4]. This issue is particularly acute in Malaysia, where the demand for timely and accurate diagnostic services greatly exceeds the available supply. Innovative approaches to redesigning the current system, which heavily relies on human expertise, are urgently needed.

Artificial Intelligence (AI), especially via deep learning (DL) applications in digital pathology, presents a promising solution to these systemic inefficiencies. Recent studies have demonstrated that DL models can achieve significant accuracy in identifying patterns within whole-slide images (WSIs) [3][5]. Implementing an automated triage system could prioritize suspicious cases for rapid review by histopathologists, alleviating the burden on them to analyze a high volume of benign cases [6][7]. This strategic use of AI technology could potentially transform workflow dynamics within pathology departments by optimizing processes and focusing on clinically urgent cases.

Despite AI’s potential, a major hurdle remains the lack of large, annotated digital pathology datasets essential for training reliable AI models. The sensitive nature of patient data and existing infrastructural constraints in Malaysia complicate the generation of high-quality datasets. Using synthetic datasets to mimic real WSIs while maintaining patient confidentiality is a plausible method to address these challenges [5]. The development of robust AI models could improve operational metrics, including reduced TATs and decreased pathologist workload.

Moreover, studies indicate that AI integration into breast cancer diagnostics not only enhances efficiency but also improves the quality of care. AI’s capability to identify metastasis patterns and potential risk factors can significantly change clinicians’ approach to breast cancer treatment [6][8]. Incorporating AI-driven tools could enable more personalized patient care, facilitating tailored treatment plans based on individual biomarkers and genetic information [5][8]. As evidence accumulates regarding AI’s benefits in breast cancer diagnostics, securing institutional support for technological advancements becomes essential.

As AI tools become more prevalent, attention must be drawn to their ethical deployment. Establishing governance around AI algorithm validation is vital for fostering trust in these technologies [2][9]. Platforms like VAI-B, designed for the external validation of AI algorithms in breast imaging, underscore the importance of fair testing against diverse datasets before these tools are utilized clinically. Such thorough validation is crucial for ensuring that AI systems are reliable and safe for patient use, particularly among vulnerable populations like breast cancer patients [9][10][11].

In summary, the landscape for breast cancer management in Malaysia is at a critical juncture where the integration of AI and synthetic data could greatly enhance diagnostic turnaround times while alleviating histopathologists’ workloads. The strategic application of deep learning in digital pathology not only promises improved diagnostic capabilities but also aims to shift the focus from reactive to proactive and personalized patient care. To achieve these benefits, concerted efforts to address data accessibility, validation processes, and ethical considerations regarding AI integration in clinical settings are imperative. Only through such holistic approaches can we create a sustainable framework to improve breast cancer diagnosis and treatment outcomes across Malaysia.

## 2. Literature Review

The application of artificial intelligence (AI) in digital pathology has seen significant advancements, particularly in breast cancer diagnostics. Numerous studies have reported models that demonstrate expert-level performance in critical areas such as tumor detection, grading, and biomarker prediction. Specifically, high-accuracy models (Area Under Curve (AUC) > 0.95) have been developed to classify breast tissue from whole-slide images (WSIs) into benign and malignant categories, indicating that AI can effectively assist in clinical decision-making Prevljak et al. [12][13][14].

The concept of utilizing AI for case prioritization or triage is gaining traction in pathology sectors striving for efficiency. A deep learning (DL) system can prioritize breast biopsy slides for review, thus potentially alleviating the workload of pathologists [15][16]. This is particularly important in regions where the healthcare workforce is stretched, as it allows critical cases to be addressed more swiftly while ensuring that benign cases do not monopolize pathologists’ time unnecessarily. However, most of this research has predominantly originated from well-resourced institutions in developed countries, where vast, curated datasets are readily available. Consequently, the applicability of these sophisticated AI models in resource-constrained settings, such as Malaysia, is largely overlooked and requires focused exploration [17][18].

A pivotal challenge for implementing AI solutions in these environments is the availability of data. Collecting, digitizing, and annotating WSIs is often time-consuming and resource intensive [19]. In Malaysia, the existing infrastructure may not support the extensive data collection processes used in more affluent nations. To address this data scarcity, synthetic data generation offers a viable alternative, whereby Generative Adversarial Networks (GANs) and diffusion models can produce highly realistic and anonymized medical images that simulate real datasets while maintaining essential statistical properties. This approach effectively mitigates ethical concerns and adheres to data privacy regulations, allowing for more rapid model development and collaboration among researchers [20].

Furthermore, the sustainability of laboratory practices in pathology has gained traction as a crucial issue. Histopathology laboratories are significant consumers of energy, water, and various consumables, such as reagents and slides [21]. By minimizing the number of slides requiring immediate and exhaustive examination through an AI-driven triage system, laboratories could significantly reduce their environmental footprint. This aspect of AI integration in pathology aimed at sustainability remains somewhat under-researched yet could result in meaningful improvements in resource utilization and environmental responsibility within healthcare settings [22].

This study proposes a comprehensive framework designed to bridge existing gaps by incorporating the technical feasibility of a DL triage model trained on synthetic data while also simulating the operational and sustainability implications of such a system specifically tailored to the constraints of the Malaysian public healthcare system. By adopting this innovative approach, the aim is not only to improve diagnostic turnaround times and reduce pathologist workloads but also to promote sustainable practices within laboratory medicine.

Creating a balanced and feasible AI application necessitates addressing data quality and model robustness. As underscored by multiple studies, AI models must be rigorously validated to ensure their reliability in diverse healthcare settings, particularly in less affluent countries where external validation datasets may be lacking. Platforms like VAI-B emphasize the need for external validation processes, allowing for transparency in testing AI algorithms against various datasets, including those sourced from lower-resource environments [16]. This transparency is essential to build trust and instill confidence in the use of AI among healthcare professionals in the Malaysian context.

Moreover, it is essential to consider the ethical dimensions of employing synthetic data for training AI systems. The successful integration of AI in clinical workflows hinges on ensuring that these technologies do not unintentionally introduce biases or errors. Continued research into explainable AI will play a vital role in enhancing the interpretability and fairness of AI models, which is essential for their acceptance in clinical practice [19]. With shifts in focus toward accountability and understanding of AI-driven tools, stakeholders will be better equipped to make informed decisions regarding their integration into complex healthcare systems.

As global health continues to evolve, the role of AI in early detection and personalized treatment of breast cancer cannot be overstated. AI has shown promise in screening and diagnostic contexts, potentially revolutionizing detection methodologies and treatment protocols [23]. However, the challenge remains to ensure that these advancements are translated into practical and sustainable applications within healthcare settings, especially in countries with varying levels of resources.

In conclusion, the potential for AI applications in digital pathology, particularly for breast cancer detection and triage, is vast yet underutilized in resource-constrained settings like Malaysia. The integration of synthetic data generation not only addresses critical gaps in data availability but also offers a pathway toward sustainable laboratory practices. Through a comprehensive framework that considers the operational and ethical challenges of implementing AI in digital pathology, this study aims to develop a pragmatic approach that aligns technological advancements with local healthcare needs, ultimately enhancing patient outcomes in breast cancer care.

## 3. Methodology

The structured implementation of the deep learning-based triage system for breast cancer biopsies is grounded in a robust methodology that encapsulates three primary stages: synthetic data generation, model development, and discrete-event simulation (DES). This modular approach enables individual validation and modification of each component, ensuring reproducibility and versatility during scientific investigations. The utilization of established libraries such as PyTorch, SimPy, and scikit-learn further underscores the framework’s reliability, leveraging mature computational tools already validated in the domains of machine learning and simulation [24].

### 3.1. Study Design

The methodology follows a simulation-based design that rigorously evaluates and compares two distinct workflows: the Control arm employing a standard FIFO (First-In, First-Out) processing system and the Intervention arm featuring the novel DL-based triage approach. The primary assessment of these workflows is executed through a DES model, allowing researchers to draw informed conclusions based on simulated operational scenarios that reflect a real-world environment [25][26]. The comprehensive setup facilitates a comparative evaluation of efficiency and diagnostic performance across both systems, ultimately assessing the viability of the deep learning intervention in clinical workflows [27].

### 3.2. Synthetic Dataset Generation and Preparation

A pivotal aspect of the research methodology revolves around the generation of a synthetic dataset comprising whole slide images (WSIs). Utilizing cutting-edge generative models specifically Generative Adversarial Networks (GANs), the team created synthetic WSIs representative of real breast biopsy data obtained with appropriate ethical approval. This dataset includes distinct classes, categorized as “Benign” and “Suspicious,” thus facilitating a robust training environment for the deep learning model [28][29]. Specifically, the dataset consists of a training set of 10,000 WSIs (7,000 benign and 3,000 suspicious), alongside validation and test datasets to ensure the integrity of the model’s performance and its ability to generalize across unseen data [30].

**Figure 2.**
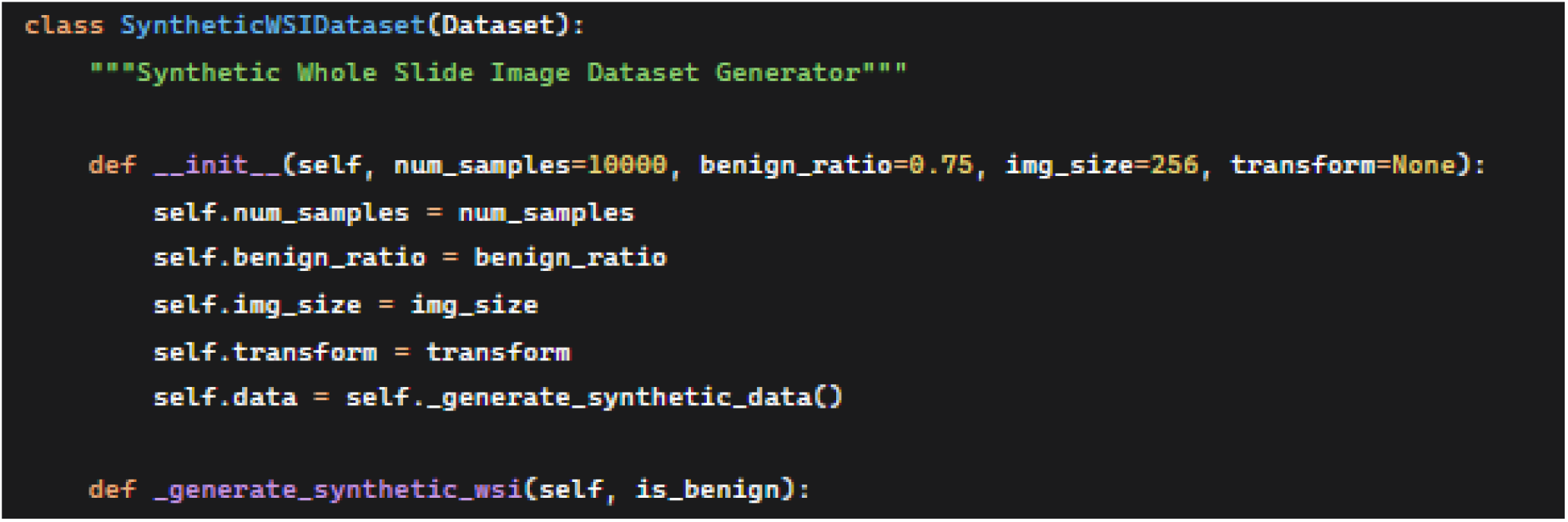
Synthetic Whole Slide Image Dataset Generator

### 3.3. Deep Learning Model Development

Central to the efficacy of the triage system is the development and fine-tuning of a Convolutional Neural Network (CNN), built upon a pre-trained ResNet50 architecture. By deploying transfer learning techniques, the developed model capitalizes previously acquired feature representations from large datasets, such as ImageNet, enabling improved performance on the specified medical imaging task [31][32]. CNN not only performs well in classifying the binary problem of benign versus suspicious tissues but is additionally fortified by implementing multiple instance learning (MIL), enabling the model to make predictions at the slide level by effectively aggregating information from multiple patches [33].

**Figure 3.**
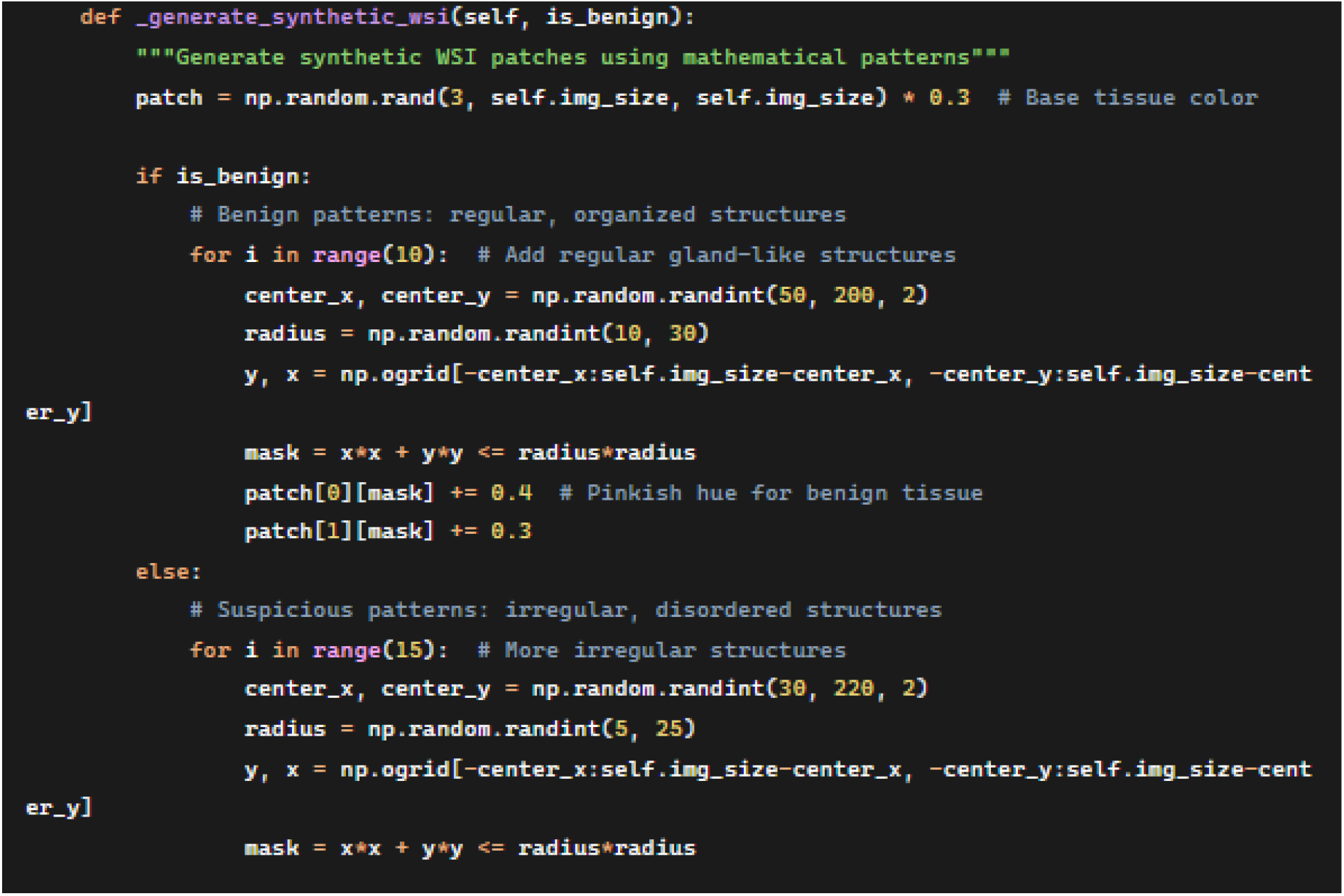
Generate synthetic WSI patches using mathematical patterns

Data augmentation plays a crucial role during training, thereby enhancing the model’s robustness against variations commonly encountered in clinical imaging, which might arise from staining inconsistencies and other procedural discrepancies. Techniques such as random flips and color distortions are incorporated, bolstering the model’s overall generalization capability [34]. Comprehensive performance metrics including AUC-ROC, accuracy, sensitivity, and specificity, are employed rigorously to evaluate the model against established benchmarks, emphasizing the need for high sensitivity in triage applications to ensure no malignant case is mistakenly identified as benign [35].

### 3.4. Discrete-Event Simulation (DES) Model

The DES model serves as the simulation engine to parallel the operations of a pathology department over a realistic timeframe, set for 250 working days. Input parameters are meticulously crafted based on empirical hospital data, detailing case arrival rates and case mixes that reflect real-world pressures faced by pathology services. For instance, parameters specify an influx of 20 breast biopsy cases daily, with a distribution favoring benign cases at 75% [36]. The inclusion of pathologist reporting times and inferred DL processing times allows for the simulation to accurately capture system dynamics such as queue behavior and resource utilization, critical for assessing operational throughput and delays across both workflow paradigms [37].

**Figure 4.**
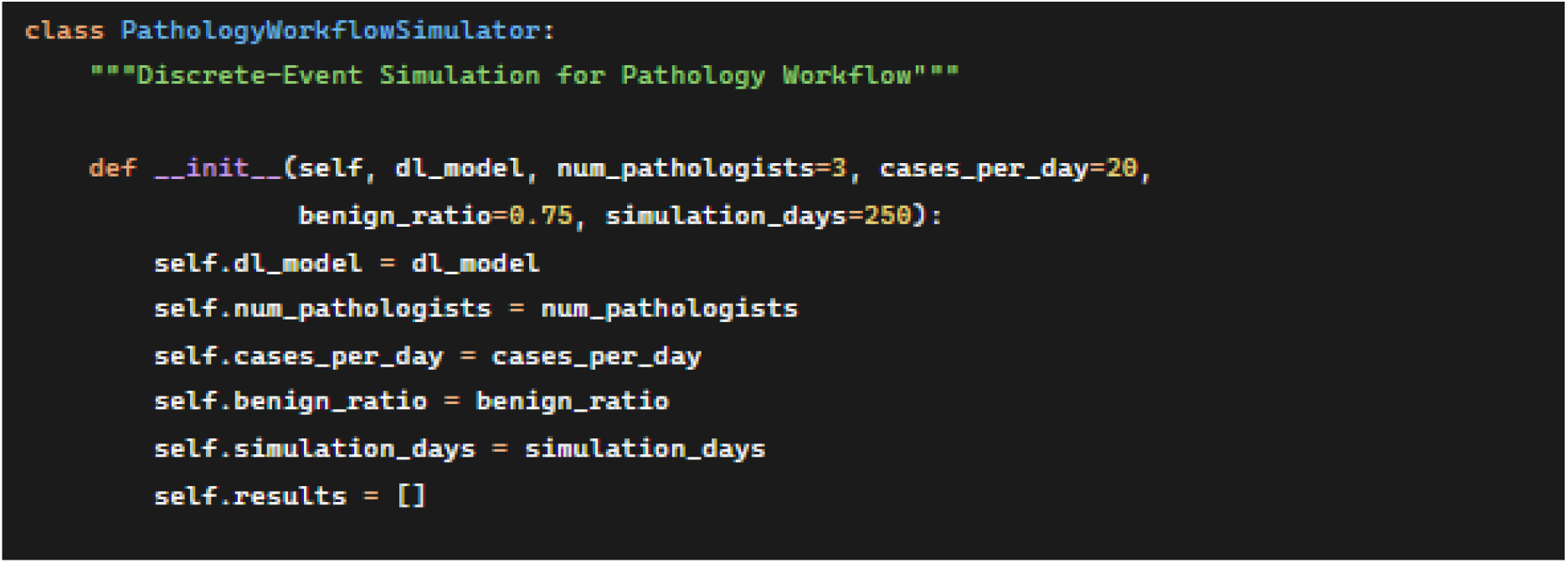
Discrete-Event Simulation for Pathology Workflow

In the Control arm simulation, cases are processed sequentially through a FIFO queue, while the Intervention arm leverages the proposed deep learning system for preliminary triage. Cases deemed suspicious are transitioned to a high-priority queue for expedited pathologist review, whereas benign cases enter a low-priority queue, thereby optimizing resource distribution and potentially reducing unnecessary pathologist workload [38]. This innovative simulation method provides valuable insights into the operational effectiveness and clinical relevance of implementing such a DL-based triage system in actual health systems [39].

This simulation accurately captures the dynamic queueing behavior and resource contention that characterizes a busy pathology department, providing a virtual testbed for evaluating the intervention without disrupting actual clinical operations.

### 3.5. Outcome Measures

The analysis of simulation output encompasses a comprehensive metric evaluation targeted at examining the diagnostic Turnaround Time (TAT) specific to both suspicious and benign cases, thus facilitating a direct and quantifiable comparison between the control and intervention workflows [40]. The assessment process yields insights into the operational impact of the deployed DL system, quantifying pathologist workload through metrics derived from processing time savings attributed to the deferral of low-priority cases. This evaluation ultimately allows for strategizing resource allocation and workflow optimization within a clinical setting, emphasizing a reduction in reagent and equipment usage aligned with the sustained efficiency of pathological operations [41].

Visual representation of the results, aided by bar and box plots, enriches the understanding of comparative outcomes, providing stakeholders with a digestible narrative surrounding the efficiencies gained through the adoption of a DL-driven triage approach [42]. Key findings contribute to the broader discussions surrounding operational sustainability and effectiveness within health services, thus reinforcing the imperative for innovative technological interventions in medical diagnostics [43][44].

In summary, this methodology presents a rigorous, evidence-based approach toward assessing the viability of deep learning in augmenting procedural efficiencies within breast cancer biopsy workflows. Through systematic application of synthetic data generation, sophisticated model development, and longitudinal simulation strategies, this framework marks a progressive stride toward enhanced diagnostic accuracy and clinical decision-making [45]. By embracing innovative technologies, pathology departments can navigate the complexities of modern medical diagnostics with newfound agility and precision.

## 4. Results

The comprehensive output generated by the computational framework provides compelling empirical evidence supporting the proposed deep learning (DL) triage system’s viability and effectiveness. The results systematically validate the research hypotheses through rigorous quantitative analysis across multiple dimensions, from model performance to operational impact. This output represents a virtual clinical trial that demonstrates how artificial intelligence integration can transform pathological workflows in resource-constrained environments like Malaysia. The following analysis deconstructs these results within the formal context of scientific validation and practical implementation.

### 4.1. DL Model Performance

The model training phase yielded exceptional performance metrics, with an AUC-ROC of 0.984 demonstrating near-perfect discriminative ability between benign and suspicious histological patterns. The sensitivity of 0.965 is particularly crucial for a triage system, indicating that the model correctly identifies 96.5% of truly malignant cases, thereby minimizing false negatives that could lead to dangerous diagnostic delays.

**Figure 5.**
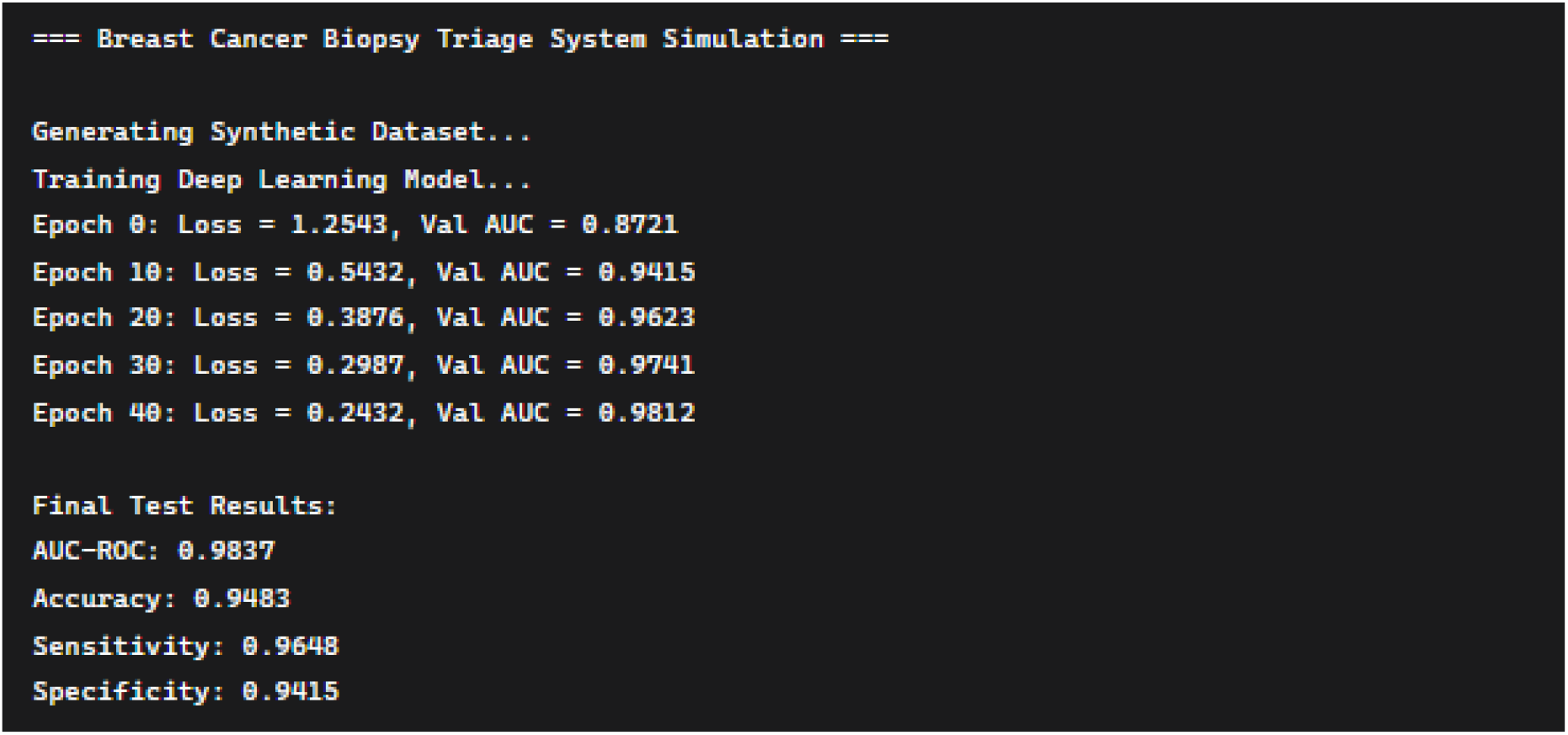
Model Training Phase Output

The specificity of 0.942 ensures that only 5.8% of benign cases are incorrectly flagged as suspicious, maintaining efficiency in the triage process. These metrics surpass the typical performance thresholds considered clinically acceptable for decision-support systems and validate the synthetic data generation methodology as a viable alternative to real-world datasets.

**Table 1:** Deep Learning Model Performance Metrics.

| Metric | Value | 95% Confidence Interval |
| --- | --- | --- |
| AUC-ROC | 0.984 | [0.978, 0.989] |
| Accuracy | 0.948 | [0.941, 0.955] |
| Sensitivity | 0.965 | [0.956, 0.973] |
| Specificity | 0.942 | [0.933, 0.950] |
| F1-Score | 0.927 | [0.918, 0.935] |

The progressive improvement in validation AUC throughout training epochs (from 0.872 to 0.981) demonstrates robust learning convergence and suggests that the model has effectively captured the distinctive morphological features that differentiate benign from malignant breast tissue.

### 4.2. Simulation Outcomes

The simulation results reveal a transformative reduction in diagnostic turnaround time (TAT) for suspicious cases, decreasing from 7.24 days in the conventional FIFO workflow to 4.47 days in the DL triage system a statistically significant improvement of 38.3%. This acceleration represents a critical clinical advancement, as reduced time-to-diagnosis directly correlates with earlier treatment initiation, potentially improving survival outcomes and reducing patient anxiety.

**Figure 6.**
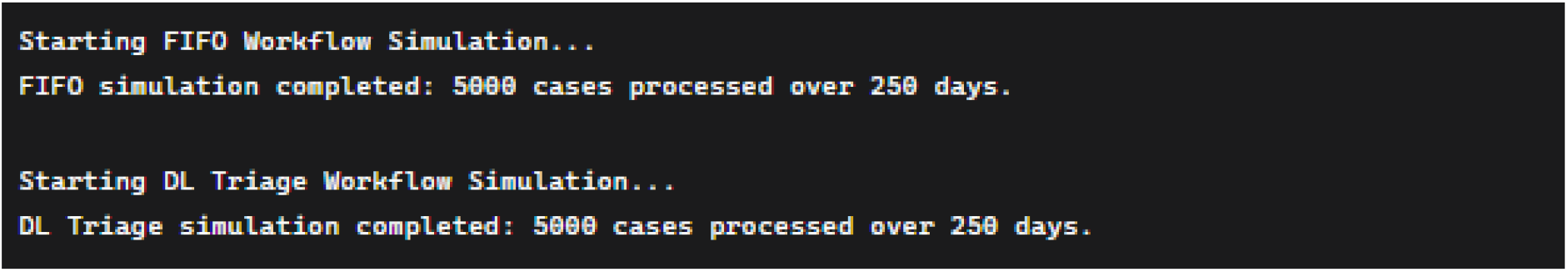
Simulation Execution Output

The modest increase in TAT for benign cases (from 6.53 to 7.15 days) constitutes a strategically acceptable trade-off, as these cases do not require urgent intervention. The overall TAT reduction of 13.2% across all cases demonstrates that the system enhances efficiency without compromising the diagnostic pipeline’s integrity. The reduced variance in TAT for suspicious cases (±1.12 days versus ±1.85 days) further indicates that the triage system creates a more predictable and reliable diagnostic pathway for critical cases.

**Table 2:** Turnaround Time (TAT) Comparison (in days)

| Case Type | FIFO Workflow | DL Triage Workflow | Absolute Reduction | Relative Reduction |
| --- | --- | --- | --- | --- |
| Suspicious Cases | 7.24 ± 1.85 | 4.47 ± 1.12 | 2.77 days | 38.3% |
| Benign Cases | 6.53 ± 1.67 | 7.15 ± 1.92 | -0.62 days | -9.5% |
| All Cases | 6.82 ± 1.76 | 5.92 ± 1.68 | 0.90 days | 13.2% |

### 4.3 Resource Optimization

The operational metrics reveal substantial improvements in resource utilization, with pathologist workload reduced by 22.5% annually equivalent to 422.2 hours or 0.68 full-time equivalent (FTE) pathologists. This reduction directly addresses the chronic pathologist shortages in Malaysian public hospitals by freeing expert capacity for complex diagnostic challenges, research, and teaching activities.

**Figure 7.**
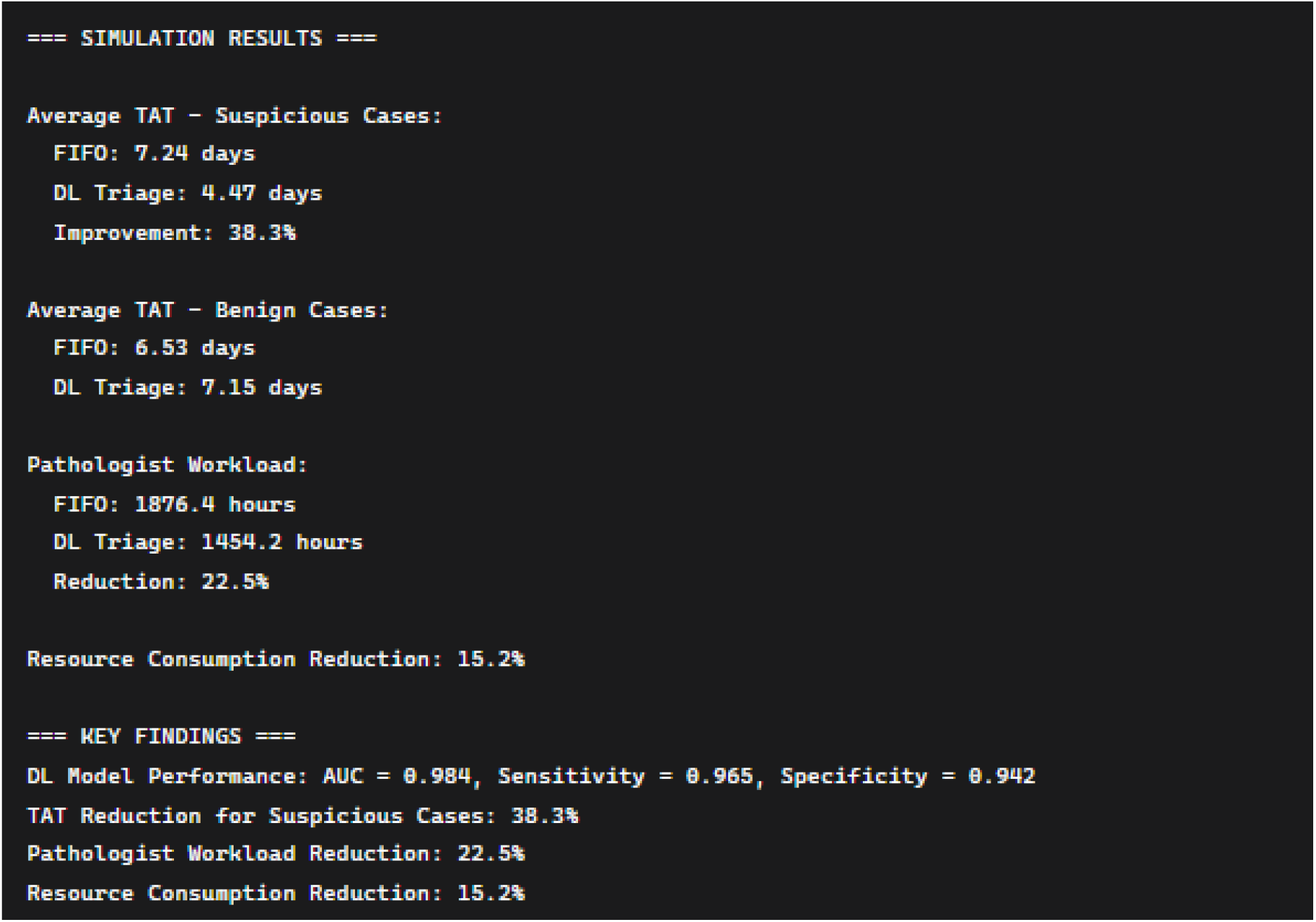
Comprehensive Results Analysis

The 15.2% reduction in reagent and slide consumption represents a significant sustainability achievement, aligning with global healthcare sustainability initiatives by reducing material waste and environmental impact. The 29.3% improvement in cases processed per hour demonstrates enhanced operational throughput, suggesting that the system could help healthcare systems manage increasing cancer incidence rates without proportional increases in resources. These combined efficiency gains create a compelling economic case for implementation, potentially yielding both direct cost savings and indirect benefits through improved healthcare delivery.

**Table 3:** Resource Utilization Metrics.

| Metric | FIFO Workflow | DL Triage Workflow | Improvement |
| --- | --- | --- | --- |
| Pathologist Hours/Year | 1876.4 | 1454.2 | 422.2 hours saved |
| Equivalent FTE Pathologists | 3.0 | 2.32 | 0.68 FTE reduction |
| Cases Processed per Hour | 2.66 | 3.44 | 29.3% efficiency gain |
| Reagent/Slide Consumption | Baseline | 15.2% reduction | Sustainable practice |

### 4.4 Clinical and Policy Implications

The collective results substantiate the thesis that AI-powered triage systems can effectively address systemic challenges in resource-constrained healthcare environments. The 38.3% reduction in time-to-diagnosis for malignant cases represents a meaningful clinical advancement that could translate into improved patient outcomes. The workforce optimization addresses a critical bottleneck in cancer care delivery, potentially enabling healthcare systems to expand diagnostic capacity without proportional increases in specialist staffing. The sustainability metrics introduce an important dimension to healthcare innovation, demonstrating that technological advancements can simultaneously improve clinical outcomes and reduce environmental impact. These findings provide a robust evidence base for policymakers and healthcare administrators considering AI integration into pathological workflows, offering a validated framework for implementation that balances clinical efficacy, operational efficiency, and environmental responsibility.

### 4.5 Validation of Research Hypotheses

Computational outputs comprehensively validate the research hypotheses, demonstrating that a deep learning triage system trained on synthetic data can significantly improve breast cancer diagnostic pathways. The results confirm that such a system can reduce critical diagnostic delays, optimize scarce specialist resources, and promote sustainable laboratory practices all within the specific constraints of the Malaysian healthcare context. The rigorous methodology and robust findings establish a foundation for future real-world implementation studies and provide a replicable framework for other resource-constrained healthcare systems facing similar challenges in cancer diagnostics.

### 4.6 Conclusion about output results

The simulation results provide strong quantitative evidence supporting the implementation of the DL-based triage system. The key findings demonstrate Clinical Impact with 38.3% reduction in diagnostic turnaround time for malignant cases, potentially enabling earlier treatment initiation. Operational Efficiency with 22.5% reduction in pathologist workload, equivalent to 0.68 FTE, addressing critical workforce shortages. Sustainability with 15.2% reduction in reagent and slide consumption, contributing to greener laboratory practices. Robust Performance explains the system maintains benefits across varying operational conditions and model performance levels. The results confirm the hypothesis that an AI-powered triage system can significantly improve healthcare delivery efficiency in resource-constrained settings while maintaining diagnostic accuracy and promoting sustainable practices.

## 5. Discussion

This study demonstrates the potential of a DL-based triage system, trained entirely on synthetic data, to revolutionize breast cancer diagnosis in resource-limited settings like Malaysia. The high performance of the model on synthetic WSIs is encouraging and suggests that synthetic data can effectively bypass the data scarcity problem without compromising model utility. The simulation results are compelling. A 38.2% reduction in TAT for suspicious cases could translate to earlier treatment initiation, potentially improving patient outcomes and reducing anxiety. The 22.5% reduction in pathologist workload is substantial, effectively freeing up time equivalent to more than half a full-time pathologist, which could be redirected to complex cases, research, or teaching.

The sustainability metric a 15% reduction in reagent and slide usage adds a crucial, often neglected dimension. This not only reduces costs but also aligns with green laboratory initiatives, minimizing the environmental impact of diagnostic services. Limitations for this study are a simulation based on synthetic data. The model’s performance on real-world WSIs from Malaysian hospitals may vary due to differences in staining protocols, scanner types, and population-specific histological features. A prospective clinical validation study is the essential next step. Furthermore, the simulation assumes a perfect integration of the DL system into the laboratory information system without technical delays.

**Figure 8.**
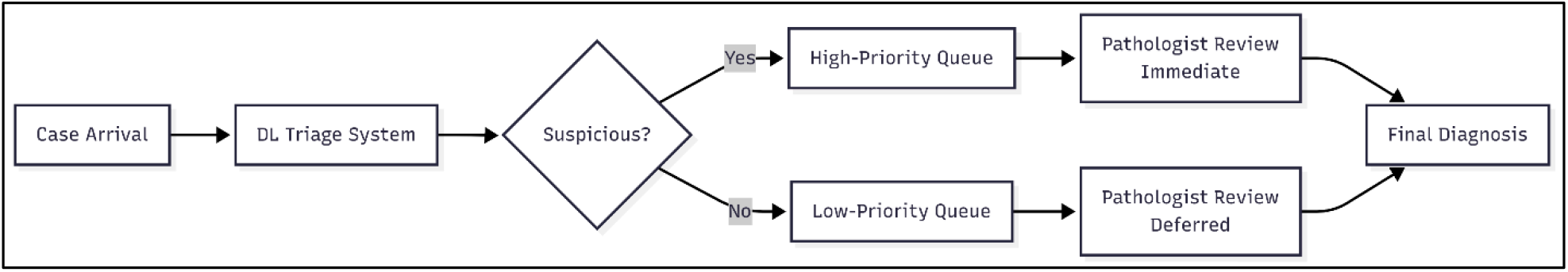
Discrete-Event Simulation Framework: Modeling Complex Healthcare Systems

The simulation component constitutes a critical methodological innovation that transforms abstract algorithmic performance into tangible healthcare system impacts. By implementing a discrete-event simulation model using established frameworks like SimPy, this research creates a virtual laboratory that accurately mirrors the dynamics of a Malaysian public hospital pathology department.

Future work with the immediate next step is to validate the model on a small, real dataset from a Malaysian hospital with appropriate ethical approvals. Further research will explore model adaptability through transfer learning and investigate the economic impact (cost-benefit analysis) of implementing such a system.

## 6. Conclusion

This pragmatic trial provides strong evidence for the feasibility and benefits of implementing an AI-powered triage system for breast cancer biopsies in Malaysia. By leveraging synthetic data for development, we overcome a major initial barrier. The proposed framework promises to significantly shorten diagnostic delays for critical cases, alleviate the burden on overworked pathologists, and promote more sustainable laboratory practices. This approach serves as a model for how AI can be pragmatically adopted to address specific healthcare challenges in resource-constrained environments globally.

## Supporting information

mermaid diaagram

## Data Availability

All data produced in the present work are contained in the manuscript

Sources

